# Current Status of Health Care Bioethics Committees in Chile: Suggestions for Improvement in Public Health Management

**DOI:** 10.1101/2024.09.23.24314157

**Authors:** Rodrigo Barra Novoa

**Affiliations:** Director, Laboratory of Innovation, Growth and Sustainability of the Andean Macro Region (Andean-Lab). PhD in Economics Science at the Camilo José Cela University (UCJC), Madrid

**Keywords:** bioethics, health care ethics committees, public health, Chile, health care management

## Abstract

This study examines the current functioning of Health Care Bioethics Committees (HBCs) in Chile, with the objective of providing recommendations for improving public health care management. Through a comprehensive literature review and a case analysis of the ‘Hospital Clínico de la Universidad de Chile’, the structure, functions and challenges of the CEA are explored. The research reveals the importance of the multidisciplinary composition of the committees, the need for continuing education in bioethics, and the relevance of the consultative role in ethical decision making. The results suggest that the effectiveness of CABs does not necessarily depend on the number of members or the length of their terms of office, but on the quality of their training and experience. It is concluded that for adequate health care management, it is essential to strengthen the role of the CEA in the ethical education of health care personnel and in the development of institutional regulations, as well as to improve their visibility and accessibility within the Chilean health care system.

## Introduction

The accelerated advance of technology and health sciences in recent decades has generated a series of complex ethical dilemmas in medical practice, posing new challenges for healthcare systems worldwide (Stahl & Coeckelbergh, 2016). In this context, the Health Care Bioethics Committees (CEA) have emerged as crucial entities to address these ethical problems in health institutions (Vergara et al., 2015).

In Chile, the implementation of CEAs has developed progressively since the beginning of the 21st century, following the international trend initiated in countries such as the United States, Spain, Brazil and Argentina (Couceiro & Beca, 2006). These committees have been established as interdisciplinary advisory bodies with the aim of analyzing and advising on ethical conflicts in health care, thus contributing to improving the quality of care and protecting patients’ rights.

Despite their growing importance, there is a significant gap in knowledge about the effective functioning of CEAs in Chile and their real impact on public health management. This study seeks to address this knowledge gap by examining in depth the structure, functioning and challenges of CEAs in the Chilean context, with the aim of proposing informed recommendations to optimize their effectiveness in the national health system in the framework of the new economy (Barra, 2021).

### Research Problem

Despite the progressive implementation of Health Care Ethics Committees (CEA) in Chile since the beginning of the 21st century, there is a significant gap in the understanding of their effective functioning and their real impact on public health care management. This lack of knowledge is manifested in several aspects:

▸ Operational effectiveness: it has not been systematically evaluated whether the current structure and composition of CEAs in Chile are optimal for addressing emerging ethical dilemmas in modern clinical practice (Beca, 2011).
▸ Impact on clinical decision-making: there is a lack of evidence on how the recommendations of CEAs influence medical decisions and patient health outcomes (Agich, 2009).
▸ Educational and policy role: The role of CEAs in ethics education of healthcare personnel and institutional policy development has not been explored in depth (Doran et al., 2016).
▸ Adaptability to new challenges: The capacity of Chilean CEAs to address emerging ethical issues related to new medical technologies and global health challenges is unknown (Jonsen, 2015).
▸ Perception and utilization: There is a lack of information on how healthcare personnel and patients perceive and use CEA services (Gaudine et al., 2011).

This study seeks to address these knowledge gaps by examining in depth the structure, operation and challenges of CEAs in the Chilean context. The goal is to propose informed recommendations to optimize their effectiveness in the national health system, thus contributing to better public health care management.

### Objectives

1. To analyze the current structure and functioning of the CEA in Chile.
2. To evaluate the effectiveness of the CEA in the resolution of ethical dilemmas in clinical practice.
3. To propose recommendations to improve public health care management through the CEA.

## Materials and Methods

### Study design

A descriptive observational qualitative study was conducted using a single case study approach. This design allowed for an in-depth analysis of the functioning of the Health Care Ethics Committee (CEA) in a specific context.

### Study site

The study analyzed the Hospital Clínico de la Universidad de Chile, located in Santiago, Chile. This hospital was selected because it is a national reference center and because of its long history of implementing an CEA.

### Inclusion criteria

▸ Documents and records of the CEA of the Clinical Hospital of the University of Chile since its implementation until 2021.
▸ Annual reports and statistics of the CEA.
▸ Ethical cases analyzed by the committee.

### Exclusion criteria

▸ Incomplete or illegible documents.
▸ Ethical cases that were not formally presented to the CEA.
▸ CEA members who participated for less than one year in the committee.

### Data collection tools

▸ Documentary review: annual reports, meeting minutes, and CEA case records were analyzed.
▸ Semi-structured interviews: Interviews were conducted with current and past members of the CEA to obtain information on its functioning and challenges.
▸ Non-participant observation: CEA meetings were attended (with proper consent) to observe its working dynamics.

### Study variables

▸ Composition of the CEA (number of members, disciplines represented).
▸ Functions performed by the CEA.
▸ Number and type of cases analyzed annually.
▸ Methodology of the committee’s work.
▸ Challenges and constraints identified by members.
▸ Perceived impact of CEA recommendations on clinical practice.

### Data analysis

A thematic analysis approach was used following the method described by Braun and Clarke (2006). Qualitative data were coded and categorized to identify emerging themes. For quantitative data (such as number of cases per year), a descriptive analysis was performed using measures of central tendency and dispersion.

### Results

The detailed analysis of the structure and functioning of CEAs in Chile, with special emphasis on the case of the Clinical Hospital of the University of Chile, revealed several significant findings:

### 1. Composition and Structure

▸ CEAs in Chile are composed of an average of 9 members, representing diverse disciplines including medicine, nursing, social work, law, and community representatives.
▸ Members’ terms of office have a standard duration of three years, with the possibility of renewal.

### 2. Functions and Activities

▸ The advisory function is the most developed, with a steady increase in the number of cases handled annually. In the case of the Hospital Clínico de la Universidad de Chile, an increase from 3 cases in 2007 to 18 cases in 2017 was observed
▸ As can be seen in Graph Nº 1, the cases consulted have been growing in number and complexity in a sustained manner (among other reasons due to the strict application of the regulatory framework on the rights of patients, the new technologies available to the Hospital, etc.). To date, there has been a change in the consultation modality, in which approximately one-third of them must be resolved in extraordinary meetings, that is, immediately and in the patient’s room.
▸ The most recurrent topics in the consultations include a) Analysis of therapeutic proportionality b) Limitation and/or adequacy of therapeutic effort c) Conflicts in medical decision-making d) Issues related to transplants and retransplants e) Maternity and perinatal ethics cases.

### 3. Working Methodology

▸ A deliberative approach to case analysis has been adopted, involving detailed fact-finding, identification of conflicting values, and proposal of concrete courses of action.
There was a growing trend towards urgent inquiries, which require responses within 24 hours.

### 4. Normative and Educational Functions

▸ Although less developed than the consultative function, significant activities were identified in these areas, including the development of internal regulations and participation in training programs for healthcare personnel.

### 5. Challenges Identified

▸ A need was noted to improve the visibility and accessibility of CEAs within healthcare institutions.

Ongoing bioethics training for committee members was identified as a crucial factor in maintaining the quality of deliberations and recommendations.

These results provide a detailed overview of the functioning of the CEA at the Hospital Clínico de la Universidad de Chile, highlighting its achievements and areas for improvement. The analysis suggests that while the CEA has succeeded in establishing itself as a relevant entity in ethical decision-making, there are significant opportunities to improve its effectiveness and impact on public health care management.

## Discussion

The results of this study suggest that CEAs in Chile have managed to establish themselves as relevant entities in ethical decision-making in the health care setting. However, there are significant areas for improvement that could enhance their impact on public health care management.

The multidisciplinary composition of the committees, which includes health professionals, ethicists and community representatives, seems to be adequate to address the complexity of modern ethical dilemmas. However, the effectiveness of CEAs does not seem to be directly related to the number of members or the length of their terms of office, but rather to the quality of their training and experience in bioethics. This is consistent with Abel’s (2006) observations on the importance of diversity and experience in the composition of committees.

The steady increase in the number of consultations attended annually suggests a growing confidence in CEAs on the part of health care personnel. However, the focus on the consultative role may be limiting the potential of these committees to more broadly influence the ethical culture of healthcare institutions. The normative function of CEAs, although less developed, shows significant potential to influence institutional policies and procedures, which could have a more lasting impact on clinical practice.

The adoption of a deliberative approach to case analysis is consistent with international best practices in bioethics (Carrese & Sugarman, 2006). However, the increasing demand for urgent consultations poses challenges to maintaining the quality and depth of ethical analysis in time-pressured situations.

The educational role of CEAs, which currently appears to be underdeveloped, represents a significant opportunity to improve the ethical awareness of healthcare personnel and thus the quality of patient care. This is particularly relevant in the context of emerging public health challenges noted by Haines et al. (2006), such as the impact of climate change on health and the ethical implications of new medical technologies.

## Conclusions

This study provides an updated and critical view of CEAs in Chile and their role in public health care management. The findings suggest that, although CEAs have managed to establish themselves as relevant entities in the Chilean health system, there are significant opportunities to improve their effectiveness and impact.

To enhance the role of the CEA as a fundamental tool for ethical and quality care management in the Chilean public health system, the following recommendations are proposed:

▸ Strengthening continuing education: Implement structured continuing education programs in bioethics for CAA members, ensuring that they are updated on the latest trends and debates in the field.
▸ Improving visibility and accessibility: Develop effective communication strategies to increase the visibility of CAAs within healthcare institutions and facilitate access to their services by healthcare personnel and patients.
▸ Expansion of regulatory and educational roles: Encourage a more active role for CAAs in institutional policy development and ethics education of healthcare personnel, drawing on their accumulated experience in resolving complex cases.
▸ Adapting to emerging challenges: Develop the capacity of CAAs to address emerging ethical issues related to new medical technologies, climate change, and other global health challenges.
▸ Systematic impact assessment: Implement monitoring and evaluation mechanisms to measure the actual impact of CEA recommendations on clinical practice and patient health outcomes.
▸ Inter-institutional collaboration: Encourage the creation of collaborative networks between CEAs from different institutions to share experiences and best practices.
▸ Inclusion of the patient perspective: Develop mechanisms to more effectively incorporate the voice and experiences of patients in the deliberations and recommendations of CEAs.

These recommendations seek not only to improve the effectiveness of existing CEAs, but also to position them as fundamental pillars in the promotion of a culture of ethics and quality in the Chilean public health system. Future studies should focus on evaluating the implementation of these recommendations and their impact on clinical practice and health outcomes.

## Limitations of the study

1. Limited generalizability: Being a single case study focused on the Hospital Clínico de la Universidad de Chile, the results may not be fully generalizable to other hospitals or contexts within the Chilean health system.
2. Temporal bias: The study is based on historical data up to 2017, which may not reflect recent changes in the structure or functioning of the CEA.
3. Lack of quantitative measurement of impact: The study did not include quantitative measurements of the direct impact of CEA recommendations on clinical outcomes.
4. Absence of patient perspective: The study did not directly include the views or experiences of patients who have been affected by CEA decisions.

## Significance statement

This study provides a critical and updated view of the functioning of the Health Care Ethics Committees in Chile, using the CEA of the Hospital Clínico de la Universidad de Chile as a case study. The research is important for several reasons:

1. It contributes to filling a gap in the literature on the practical functioning of CEAs in the Chilean context.
2. It offers valuable insights into the challenges and opportunities for improving the effectiveness of CEAs in public health care management.
3. Provides evidence-based recommendations to strengthen the role of CAAs in promoting an ethical culture in the health system.
4. Emphasizes the importance of continuing education in bioethics and the need to adapt CEAs to emerging challenges in public health.
5. It lays the groundwork for future research on the impact of CEA on clinical practice and patient health outcomes.

This research is crucial for health policy makers, hospital administrators and health professionals interested in improving the ethical quality of health care in Chile and potentially in other Latin American countries.

## Data Availability

All data produced in the present work are contained in the manuscript

## Author contributions

The research process began with an exhaustive review of the existing literature on Health Care Ethics Committees in Chile and internationally. This theoretical basis was complemented with a comprehensive analysis of the work of the Clinical Hospital of the University of Chile.

The study methodology was carefully designed to capture the complexity of the functioning of the CEA, using a single case study approach that allowed for a detailed and contextualized analysis. Data analysis was conducted using thematic analysis techniques, which allowed for the identification of patterns and emerging themes in the large amount of information collected.

The writing of the manuscript was carried out with a critical and reflective approach, seeking not only to describe the current situation of CEA, but also to provide informed recommendations to improve its effectiveness in the Chilean health system. The writing process involved multiple revisions and refinements to ensure the clarity and academic rigor of the text.

The study benefited from the researcher’s previous experience in projects related to the new economy and digital health in Chile, which allowed the findings to be contextualized within a broader framework of health system transformation. Finally, this study represents a significant contribution to the field of bioethics and health management in Chile, offering a valuable perspective on the functioning of Health Care Ethics Committees and their potential to improve the quality of health care.

---

**Graph 1.**
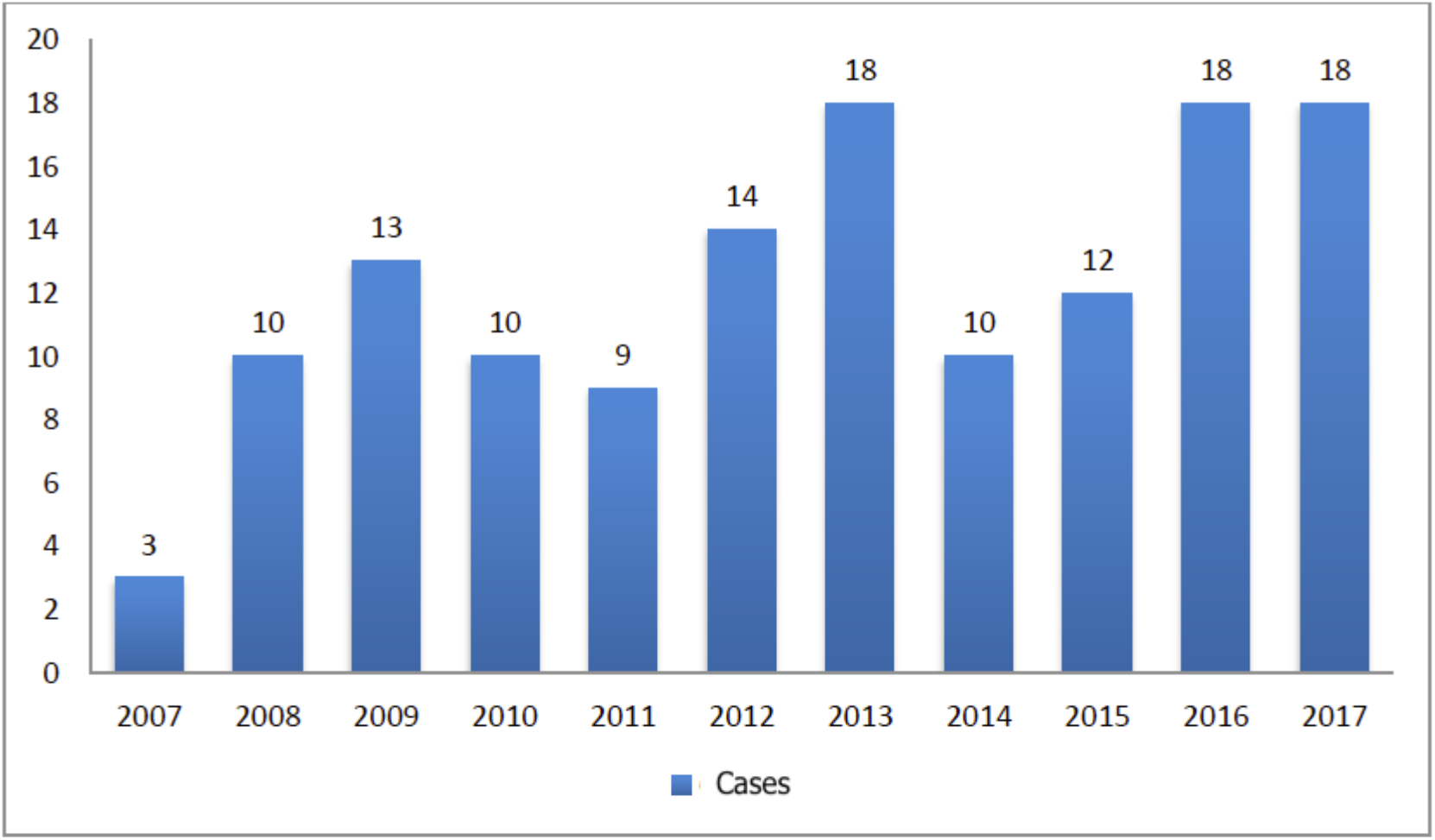
Variation of Cases 2007 - 2017. Source: Healthcare Ethics Committee Hospital Clínico Universidad de Chile

